# “CD4^+^ T Cells drive Inflammation in Type 2 Diabetes Mellitus via the TNF-α/STAT-3 Signaling Pathway”

**DOI:** 10.1101/2024.11.20.24317638

**Authors:** Shubham K Shaw, Soumya Sengupta, Rohila Jha, Chandrasekhar Pattanaik, Harapriya Behera, Prakash K Barik, Dayanidhi Meher, Rajlaxmi Sarangi, Satish Devadas

## Abstract

**Objectives:** To establish adaptive immune cells specifically T helpers as mediators of meta-inflammation in Type 2 Diabetes Mellitus, correlate biochemical and immunological parameters and delineate the specific signaling proteins responsible for it.

**Research Design and Methods:** 100 T2DM patients with no other clinical disease, autoimmunity or infection were recruited and analyzed for their biochemical and immune parameters. Bioplexing and flow cytometry was employed to analyse total and cell specific protein secretion respectively. *Ex-vivo* inhibition studies were performed using targeted monoclonal antibodies or small molecule STAT inhibitors.

**Results:** CD4^+^ T-cells were found to be the primary source for meta-inflammation in T2DM patients with multiple pro-inflammatory cytokines and antibody isotypes. TNF-α acting through STAT-3 was shown as the primary pathway implicating meta-inflammation through CD4^+^ T-cells, wherein inhibitor studies revealed subtle pathways differences between TNF-α or STAT-3 inhibition.

**Conclusions:** Our result suggests that chronic meta-inflammation with a dysregulated biochemical profile have severe implications on immune function. Additionally, TNF-α and STAT-3 inhibition are good therapeutic targets for better T2MD treatment in ameliorating meta-inflammation.

## Introduction

Type 2 diabetes mellitus (T2DM) has been increasingly recognized as a multi organ metabolic and inflammatory disease characterized by persistent hyperglycaemia, chronic low-grade inflammation and dysfunction involving the immune system. Meta-inflammation in T2DM is well-documented and associated with diabetes-related complications such as chronic kidney disease (CKD), retinopathy, cardiomyopathy, hepatopathy, neuropathy, diabetic foot ulcerations (DFU) etc [1, 2]. Despite this generalised understanding of covert inflammation involved in multi organ damage in T2DM, the exact and or specific sources including the immune system driving meta inflammation in T2DM remain poorly defined.

Additionally, the lack of correlation between glycated haemoglobin (HbA1c) levels, and the aberrant inflammatory immune response negatively impacts effective therapeutic interventions and has major organopathies implicated. Most importantly, the presumption of good health with passable carbohydrate, lipid, general organ health parameters, etc. with higher than normal Hb1Ac has serious immunological retributions when covert meta inflammation is not addressed [3]. Chronic T2DM has immune implications bordering on mild to severe depending on glucose levels and reflects more severely during an infection or injury [4-6]. Thus, identifying link between glucose dysregulation, immune activation, inflammation etc. could aid in early diagnosis, prognosis, and therapeutic approach for T2DM.

We have previously shown that T2DM patients with COVID-19 exhibit elevated cytokine levels, higher amounts of non-protective antibodies, and an altered immune cell profile, all correlating with meta-inflammation [7]. Both the innate and adaptive immune systems are known to contribute to T2DM-related inflammation [7, 8]. Recently, the role of adaptive immunity specifically T lymphocytes in the pathogenesis of T2DM has been highlighted [9] along with dysregulated immune profiles in T2DM. CD4^+^ T helper cells with elevated pro-inflammatory markers have been shown to infiltrate both visceral and subcutaneous adipose tissue [10].

Additionally, studies using obese mouse models have illustrated the involvement of T cells in adipose tissue inflammation [11], suggesting the need for further investigation into T-cell roles’ in T2DM patients.

Research on immune cell involvement in T2DM has largely focused on innate immune cells, such as macrophages, dendritic cells, and neutrophils [12-14]. However, their short lifespan and lack of memory function limit their role in sustaining long-term low-grade inflammation. In contrast, adaptive immune cells, particularly T cells, may provide a more comprehensive view of inflammatory processes underlying T2DM. while presently the specific contributions of T helper cells to T2DM exacerbation are not yet fully understood. Chronic meta-inflammation in T2DM has been linked to certain immune mediators, such as TNF-α, IL-6, and IL-1β and signalling pathways such as NF-κb and NLRP3 inflammasome. However, other cytokines—including GM-CSF, IL-17, and IFN-γ and signaling pathways, such as STATs and MAPKs, have not been extensively studied, though they may offer valuable insights into disease progression and therapeutic responses.

In this study, we demonstrate that TNF-α secreted by T helper cells may be associated and be causative for the elevated levels of basal inflammation and antigen activated response observed in T2DM patients. Our findings conclusively demonstrate that both naïve and activated T helper cells from T2DM patients display an aberrant inflammatory phenotype than healthy controls. Additionally, we present evidence for sustained and elevated levels of inflammatory cytokines, antibody isotypes, and immune cells in T2DM patients, correlating these with HbA1c.

Previous studies from our laboratory and the present study clearly demonstrate that elevated cytokine levels drive plasma B cell differentiation that secrete unwarranted anti body isotypes. Thus both arms of the adaptive immune system along with an altered innate immune arms ferment basal inflammation eventually leading to organ damage. We, further characterize immune dysregulation in T2DM, with a focus on T helper cell compartments and their subsets, to elucidate the immune mechanisms underlying this disease. We also identified STAT3 signalling pathway as downstream pathway playing a central role in mediating inflammation.

## Research Design and Methods

### Clinical Details of T2D patients

Clinical parameters such as glycated haemoglobin levels (HbA1c), fasting and average glucose levels, CRP, Lipid profile etc. was provided from Department of Biochemistry, Central Laboratory (NABL accredited), KIMS Hospital. 100 DM confirmed subjects (HbA1c >6) were recruited from KIMS Hospital, Bhubaneswar, Odisha with 40 age and sex-matched healthy subjects devoid of any underlying disease, autoimmunity or infection. All the T2DM participants recruited were on anti-diabetic medication and were free of co-morbidities or other known disease or infections. Clinical parameters such as glycated haemoglobin levels (HbA1c), fasting and average glucose levels, CRP, Lipid profile etc. was provided by KIMS Hospital medical personnel. Signed Consent forms were obtained from all participants in the study.

### Plasma cytokines, chemokines, soluble receptors, growth factors and antibody isotypes detection assay

Neat plasma derived from T2DM patients and healthy controls were run in duplicates to measure 46 different analytes including cytokines, chemokines, soluble receptors, and growth factors using a Human ProcartaPlex Mix & Match 46-Plex kit (Cat. No PPX-46-MX324DE, Invitrogen, Vienna, Austria), based on manufacturer’s instructions. Another set of neat plasma from T2DM patients and healthy volunteers, were analysed by ProcartaPlex Human Antibody Isotyping Panels (Cat. No EPX070-10818-901, Invitrogen, Vienna, Austria). The samples were acquired in a Bio-Plex 200 system and analyte concentrations were calculated using Bio-Plex manager software with a five-parameter (5PL) curve-fitting algorithm applied for standard curve calculation [15, 16].

### PBMC isolation, Surface and intracellular staining for Flow Cytometry

PBMCs was isolated via density gradient centrifugation from T2DM patients and healthy volunteers. Isolated cells were then activated using PMA (20μg/ml) and Ionomycin (1μg/ml) for 8 hours. Brefeldin (5μg/ml) and Monensin (2μmol) was added during the last 4 hours and dead cells were excluded using a Zombie Fixable NIR or Aqua Dye Kit (Bio Legend, San Diego, CA, USA). Intracellular cytokine staining was performed with fluorochrome tagged surface markers, cytokines, and transcription factors, using a Cytofix/Cytoperm Fixation/Permeabilization Solution Kit (BD Biosciences, San Jose, CA, USA). In contrast, transcription factor staining was performed using a FOXP3 staining buffer set (eBioscience, San Diego, CA, USA), according to the manufacturer’s protocol. The details of the gating strategy are given in the supplementary figures.

### Intracellular Phospho-Protein Staining

For pSTAT expression, cells were stimulated with PMA (20 ng/ml) and Ionomycin (1 mg/ml) for 6 hours, fixed with 2% formaldehyde for 30 minutes at RT, permeabilized with 1% Triton-X and 90% Methanol, washed with eBiosciences Perm wash buffer. Cells were then stained with fluorochrome-labeled phosphor STAT antibodies for 30 minutes, washed, and acquired in BD LSR Fortessa.

### *Ex-vivo* Inhibitor studies

For inhibitor studies, T2DM PBMCs were stimulated with PMA (20 ng/ml) and Ionomycin (1 μg/ml) for 16 hours in the presence of cytokine or pSTAT inhibitors, Brefeldin (5μg/ml) and Monensin (2μmol) was added during last 12 hours of stimulation. After 16 hours cells were washed and dead cells were excluded using a Zombie Fixable Aqua Dye Kit (Bio Legend, San Diego, CA, USA). Intracellular staining was performed as described previously and data acquired on a BD, LSR Fortessa

### Confocal Microscopy

For pSTATs nuclear localization studies, isolated CD4^+^ T cells were stimulated with PMA/Ionomycin for 6 hours and stained for pSTAT1 and pSTAT3 expression based on the above-mentioned protocol. Subsequently, the cells were smeared on pre-coated Poly-L-Lysine slides with anti-fade and DAPI. TCS SP5 Leica confocal microscope was used to visualize pSTAT staining using 488nm and 640nm lasers [17].

### Design of Statistical Analysis

Non-parametric tests (such as paired t-tests, Mann-Whitney tests, Kruskal Wallis tests, and Two-way ANOVA) and were performed using GraphPad Prism 8.0.1.

## Results

Our findings **(Table 1)** report a dysregulated biochemical profile in T2DM patients. Among the 38 (38%) females and 62 (62%) males DM patients, the mean age was 53.1 ± 10.4 years, HbA1c levels are slightly elevated in T2DM patients, with mean HbA1c reaching up to 7.6±1.5. The estimation of blood glucose levels revealed mean FPG to be 141±71 mg/dl, mean PPPG was 213±77 mg/dl, whereas mean random plasma glucose was 128±26 mg/dl and average blood glucose was found to be 175±44 mg/dl. In the lipid profile data, mean total cholesterol was 180± 45 mg/dl, LDL cholesterol was 105 mg/dl, HDL cholesterol was 46±20 mg/dl and Triglycerides was 192±100 mg/dl, while mean cholesterol HDL ratio was 4.4±1.9. In the kidney profile data mean serum Creatine was 1.22±1.21 mg/dl, Sodium was 137.05±4.45 mmol/L, Albumin: was 4.11±0.38 g/dl, and Globulin was 3.1±0.6 g/dl.

**Table 1.** Baseline demographics and clinical characteristics of healthy control subjects and T2DM patients: HC, HC, Healthy controls; T2DM, Type 2 Diabetes mellitus; Hb1AC, glycated haemoglobin; FPG, Fasting Plasma Glucose; PPPG, Postprandial Plasma Glucose; HDL, High-density lipoprotein; LDL, low-density lipoprotein; CRP,Creactive protein; SGOT, serum glutamic-oxaloacetic transaminase; SGPT, serum glutamic-pyruvic transaminase; N.R, Normal range Age values are represented as %. Other clinical parameters are represented as Mean ± SD

| Characteristics | HC (40) | T2DM (100) |
| --- | --- | --- |
| Age | 35±8 | 53.13±10.46 |
| Female | 45% | 38% |
| Male | 55% | 62% |
| HbA1c (<5.7) | <5.5 | 7.6±1.5 |
| Random plasma Glucose (mg/dl) (70-140) | N.R | 128±26 |
| FPG (mg/dl) (70-100) | N.R | 147±71 |
| PPPG (mg/dl) (70-140) | N.R | 213±77 |
| Avg. blood Glucose | N.R | 175±44 |
| Total cholesterol (mg/dl) (<200) | N.R | 180±45 |
| HDL (mg/dl) (>40) | N.R | 46±20 |
| LDL (mg/dl) (<130) | N.R | 97±34 |
| HDL ratio | N.R | 4.4±1.9 |
| Triglycerides (mg/dl) (<150) | N.R | 192±100 |
| Serum Creatine (mg/dl) (0.5 - 1.3) | N.R | 1.22±1.21 |
| Serum Sodium (mg/dl) (136.0-146.0) | N.R | 137.05±4.45 |
| Serum Globulin | N.R | 3.1±0.6 |
| Serum Albumin (mg/dl) (3.5-5.0) | N.R | 4.11±0.38 |
| SGPT (U/L) (5.0-40.0) | N.R | 31±23 |
| SGOT (U/L) (0.0-40.0) | N.R | 33±32 |
| Serum Alkaline Phosphatase (U/L) (35-129) | N.R | 95±33 |
| **Medications:**<br>Insulin: 11%<br>Oral Hypoglycaemics: 89%<br>(Metformin, Glimeperide, Gliptins; Vilda, Sita, Lina, Voglibose, Dapagliflozin, Empagliflozin) |  |  |

In liver function data, mean SGPT was 31±23 U/L, SGOT was 33±32 U/L and Serum Alkaline Phosphatase was 95±33 U/L. 11% of T2DM patients were on insulin, while 89% were on oral hypoglycaemics (Metformin, Glimepiride, Gliptins, Vildagliptin, Sitagliptin, Linagliptin, Voglibose, Dapagliflozin, Empagliflozin). Clinical characteristics of T2DM patients showed higher lipid and dysregulated kidney profiles as compared to healthy controls.

The clinical and biochemical profile of T2DM patients displayed higher than control Carbohydrate and Lipid levels suggesting dysregulation and thus we examined for possible co-relation to chronic low-grade meta-inflammation, by measuring total circulating inflammatory markers including cytokines, chemokines and antibody isotypes. Amongst 46 different analytes we measured in neat plasma samples, T2DM patients displayed significantly higher inflammatory proteins including antibody isotypes than healthy controls. We observed elevated levels of 27 plasma proteins including 18 cytokines, 3 chemokines, 4 Growth factors and 2 soluble receptors and labelled them as high, medium, and low expressers (Fig- 1. A) based on comparison with healthy controls. Among the high producers we observed statistically significant levels of IL-17, IL-18, IL-6, IL-3, IL-10, HGF (p values <0.0001), IL-1β (p value = 0.0002) and IL-4 (p value = 0.0007). Among the medium producers we observed significant levels of TNF-α (p value = 0.0062), VEGF-D (p value = 0.0034), GMCSF and MIP-1α (p value = 0.0013), GITR (p value = 0.0024), GCSF (p value = 0.0054), CD62E (p value = 0.0019), TNFRI (p value = 0.0035), TPO (p value = 0.0047), Insulin (p value = 0.0036), IL-5 (p value = 0.0011), IFN-α (p value = 0.0044), IL-2 and EOTAXIN (P value = 0.0033), IL-15 (p value = 0.0095), and IFN-γ (p value = 0.0016). We further examined 7 different antibody isotypes in T2DM patients to validate if increased cytokines levels for a sustained period of time between 45-60 days, could lead to an increase in circulating antibody Isotypes, as suggested by several reports [7, 18]. Not surprisingly, all 7 antibody isotypes analyzed, displayed a significant statistical increase in T2DM patients when compared with healthy controls including IgG1, IgG3, IgG4, IgA, IgM (with p value < 0.0001), IgE (p value = 0.0025) and IgG2 (p value = 0.0062) (Fig- 1. B).

**Figure- 1.**
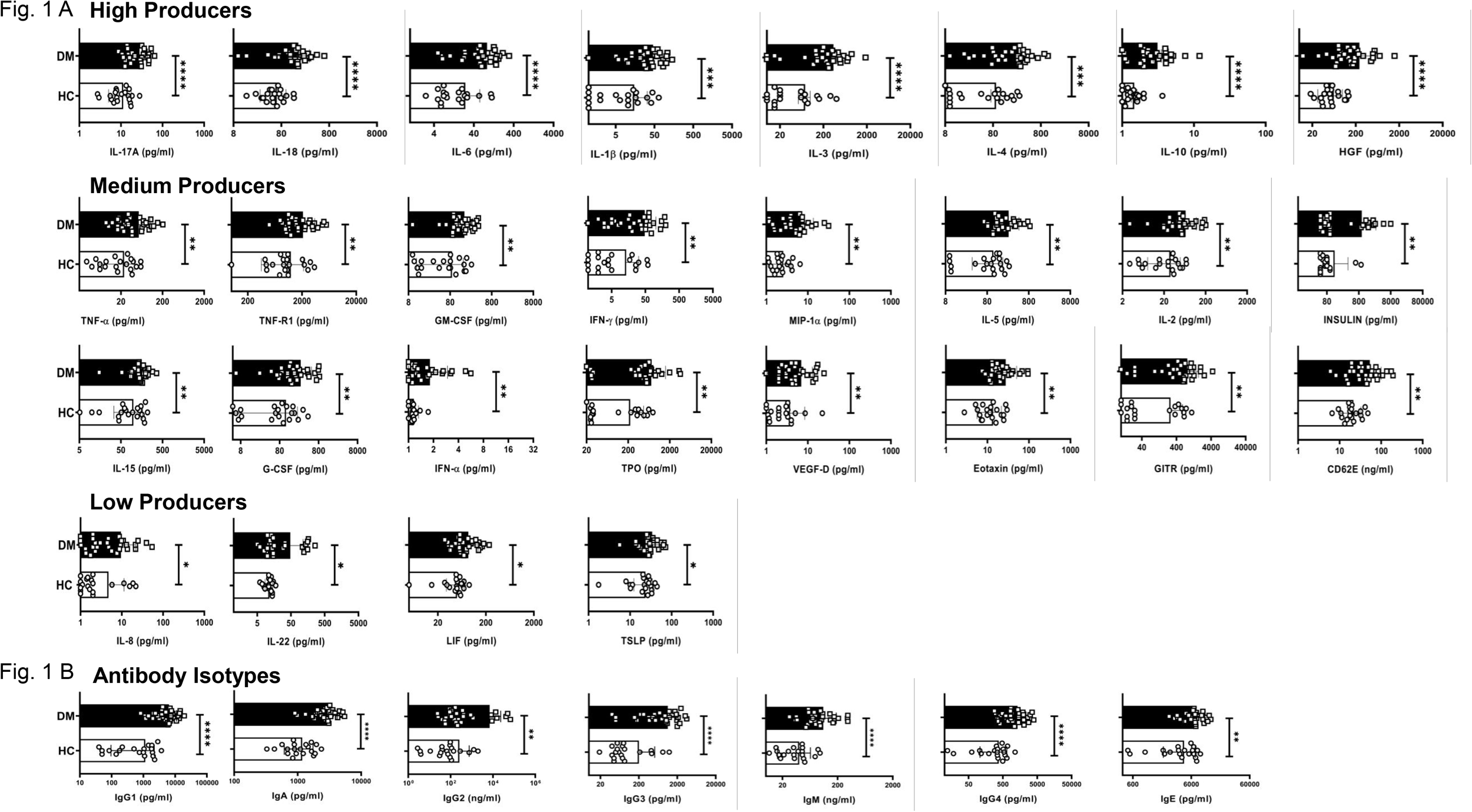
Elevated expression levels of plasma cytokines and Antibody Iso types: Representative figure of differentialy augmented levels (high, medium and low producers) of the cytokines, chemokines, growth factors and soluble receptors in plasma isolated from peripheral blood of T2DM patients (n=30) vs. healthy controls (n=20). IL-17A, IL-18, IL-6, IL-1b, IL-3, IL-4, IL-10 and HGF were highly elevated in T2DM patients when compared with healthy controls and designated as high producers. TNFa, TNF-R1, GM-CSF, IFN-g, MIP-1a, IL-5, IL-2, Insulin, IL-15, G-CSF, IFN-a, TPO,VEGF-D, Eotaxin, GITR, CD62B were moderately elevated in T2DM patients when compared to healthy controls and designated as medium producers. IL-6, IL-22, LIF and TSLP were slightly elevated in T2DM patients when compared to healthy controls and designated as low producers **(A)**. Increased expression of the circulating antibodies such as IgG1, IgA, IgG2, IgG3, IgM, IgG4 and IgE in plasma isolated from peripheral blood of T2DM patients (n=30) vs. healthy controls (n=20) **(B)**. Error bar in the above bar diagrams indicates SD. Mann– Whitney U Test was performed to compare between the two groups; p < 0.05 was considered statistically significant (*); p < 0.01 was considered to be very significant (**); p < 0.001 was considered to be highly significant (***); p < 0.0001 was considered extremely significant (****), and ns means non-significant.

In our next step, the cellular source of inflammatory cytokines in T2DM patients were analyzed with immune cell surface markers and the inflammatory cytokines to establish metabolic dysregulation, inflammatory cytokine bias and aberrant immune cells. Amongst 30 T2DM patient samples analyzed, all displayed statistically significant higher basal levels of pro-inflammatory cytokines, such as IL-17 (p value = 0.0124), TNF-α (p value < 0.0001), and GM-CSF (p value = 0.0047), secreted by CD4^+^ T cells even in their “resting” state (Fig- 2. A, B) apart from slightly higher secretion from CD8^+^ and CD14^+^ cells and were completely absent in healthy controls.

**Figure- 2.**
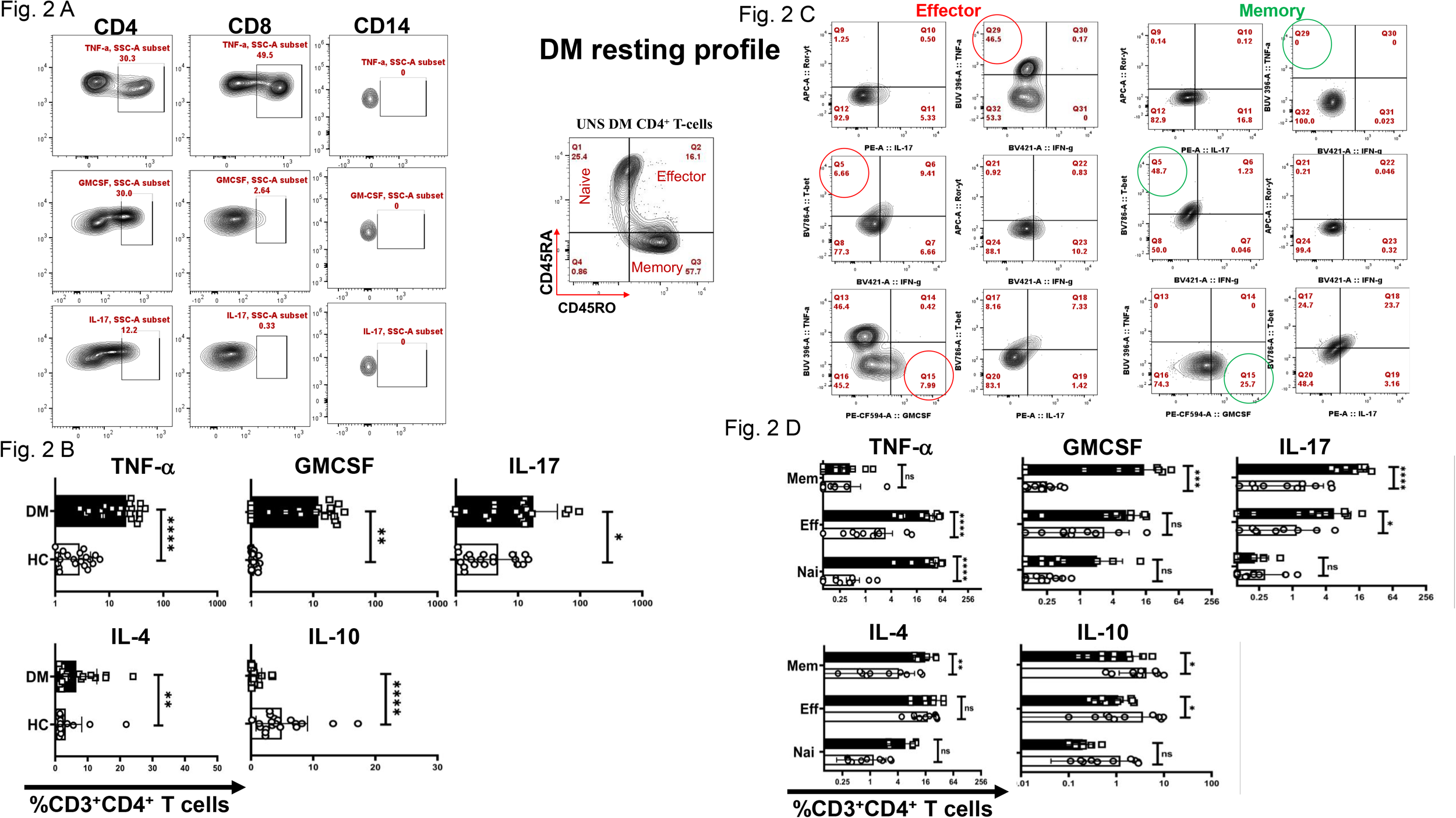
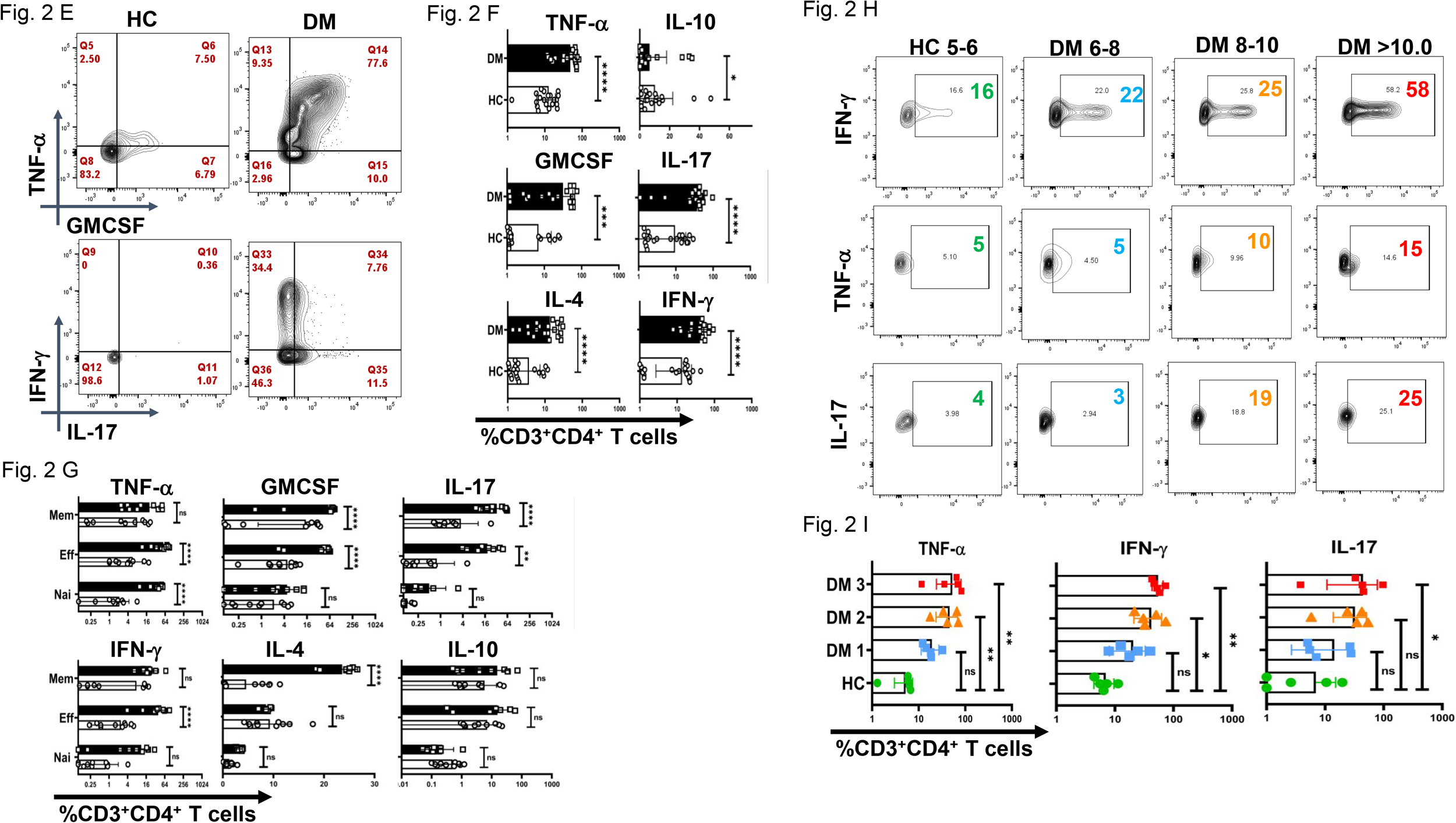
Basal expression of cytokine levels from CD4^+^ T-cells and its compartments. “Resting” state CD4+ cells of T2DM patients exhibit significantly higher basal levels of pro-inflammatory cytokines, such as IL-17, TNF-a, and GM-CSF followed by higher secretion from CD8^+^ and CD14^+^ cells as opposed to healthy controls as represented in flow cytometry plots and analyszed by gating on live CD3^+^CD4^+^ T-cells in PBMCs **(A)** and statistical graphical plots represents cytokine production from CD4^+^ T-cells **(B)**. Additionally, T-cell compartment analysis was done by gating on live population of CD3^+^CD4^+^CD45RA^+^CD45RO^-^ for naive T-cells, CD3^+^CD4^+^CD45RA^+^CD45RO^+^ for effector T-cells and CD3^+^CD4^+^CD45RA^-^CD45RO^+^ for memory. Aamongst pro-inflammatory cytokines, effector and naive resting CD4^+^ T cells secrete elevated levels of TNF-a and memory compartment predominantly secrete GM-CSF and IL-17. Anti-inflammatory cytokines such as IL-4 expression is significantly higher and from the memory compartment, IL-10 expression is lower in both effector and memory compartments compared to HCs are represented as flowcytometry plots **(C)** and cumulative graphical representation **(D). “**Activated” CD4^+^ T cells of T2DM shows higher expression of multiple pro-inflammatory cytokines phenotype as compared to healthy controls represented by flow cytometry analysis by the same gating applied in resting T-cells **(E)** and cumulative graphical representation showing higher expression of inflammatory cytokines and anti-inflammatory cytokines such as TNF-a, GMCSF, IL-17, IL-4 and IL-10 as compared to healthy controls **(F)**. T-cell compartment analysis of **“**Activated” CD4^+^ T cells of T2DM shows higher expression of pro-inflammatory and anti-inflammatory cytokines such as IL-4 expression is significantly from the memory compartment as compared to healthy controls represented by graphical plots **(G)**. Positive correlation is observed between HbA1c levels and pro-inflammatory cytokines in 3 different groups of T2DM patients with increasing levels of HbA1c such as DM1 (6.0-7.5), DM2 (7.5-9), and DM3 (<9) **(H)**.

Further investigation into specific T helper cell compartments revealed that a significant majority of Effector CD4^+^ T cells secreted TNF-α while GM-CSF and IL-17 secretion was predominantly from the memory compartment (Fig- 2. C, D).

Interestingly, even the naïve compartment secreted TNF-α establishing an activated naïve T cells in the “resting” state.

We also examined anti-inflammatory cytokine expression, including IL-4, IL-5, and IL-10. Notably, CD4^+^ T cells in T2DM patients secreted significantly higher levels of IL-4 (p value = 0.0026), predominantly from the memory compartment, while IL-10 (p value < 0.0001) expression was markedly lower in both effector and memory compartments compared to HCs. At the same time, IL-5 levels did not exhibit statistical significance between T2DM patients and HCs.

Activated T2DM CD4^+^ T cells exhibited significantly higher levels of pro-inflammatory cytokines, including TNF-α, IL-17, and IFN-γ (with p values < 0.0001), GM-CSF (p value = 0.0008), compared to HCs (Fig- 2. E, F). We report an aberrant CD4^+^ T cell phenotype in T2DM, characterized by the secretion of multiple pro-inflammatory cytokines, with a subset of double-positive TNF-α and GM-CSF cells that also secreted IFN-γ and IL-17. This aberrant T-cell phenotype was predominantly observed in both effector and memory compartments. Upon activation, the expression levels of IL-4 were consistently higher in the memory compartment with IL-10 expression being consistently lower from healthy controls.

Finally, we investigated correlations between biochemical parameters and immune profiles. Also, a significant group of T2DM population displayed a potential correlation between a dysregulated biochemical profile specifically elevated HbA1c levels and increased pro-inflammatory cytokine signatures, including IFN-γ (p value = 0.0020), TNF-α, (p value = 0.0042), and IL-17 (p value = 0.0365) in 3 different groups of T2DM patients with increasing levels of HbA1c such as DM1 (6.0-7.5), DM2 (7.5-9), and DM3 (<9) (Fig- 2. H, I).

Next, our objective was to investigate the signaling proteins potentially responsible for the elevated TNF-α expression, given that TNF-α was the primary pro-inflammatory cytokine expressed in T2DM patients, predominantly through the effector compartment. To address this, we examined the phosphorylation status of STAT proteins, including pSTAT1, pSTAT3, pSTAT4, pSTAT5, and pSTAT6, following activation with PMA/Ionomycin. Our findings revealed significantly higher levels of phosphorylated pSTAT1 (p value = 0.0068), and pSTAT3 (p value < 0.0001) in CD4^+^ T cells from T2DM patients compared to healthy controls (Fig- 3. B, C). Confocal microscopy further confirmed this result, showing nuclear co-localization of pSTAT1 and pSTAT3 in CD4^+^ T cells from T2DM patients but not in healthy controls (Fig- 3. A). Phosphorylation of other STAT proteins did not show significant differences between the two groups.

**Figure- 3.**
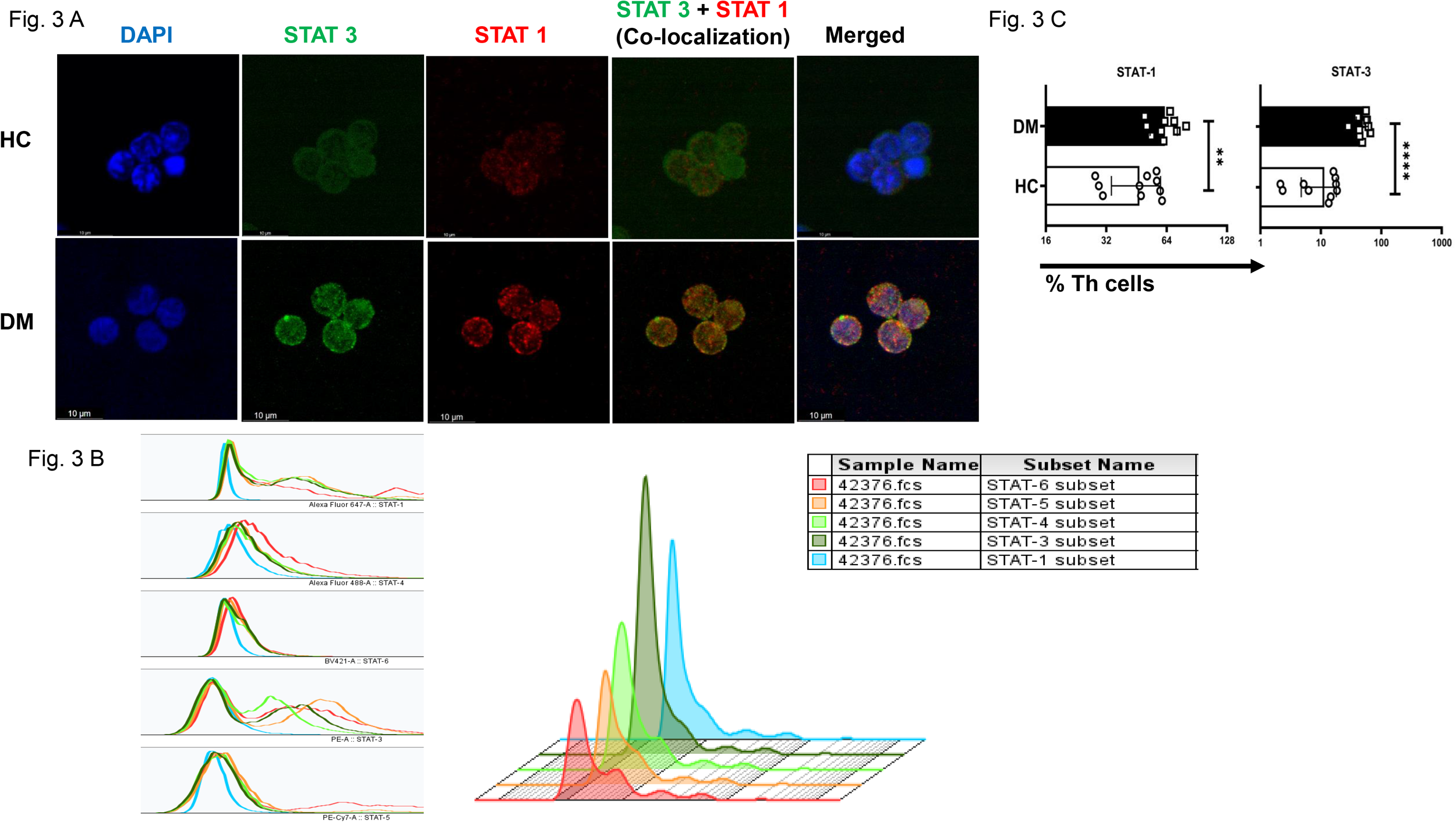
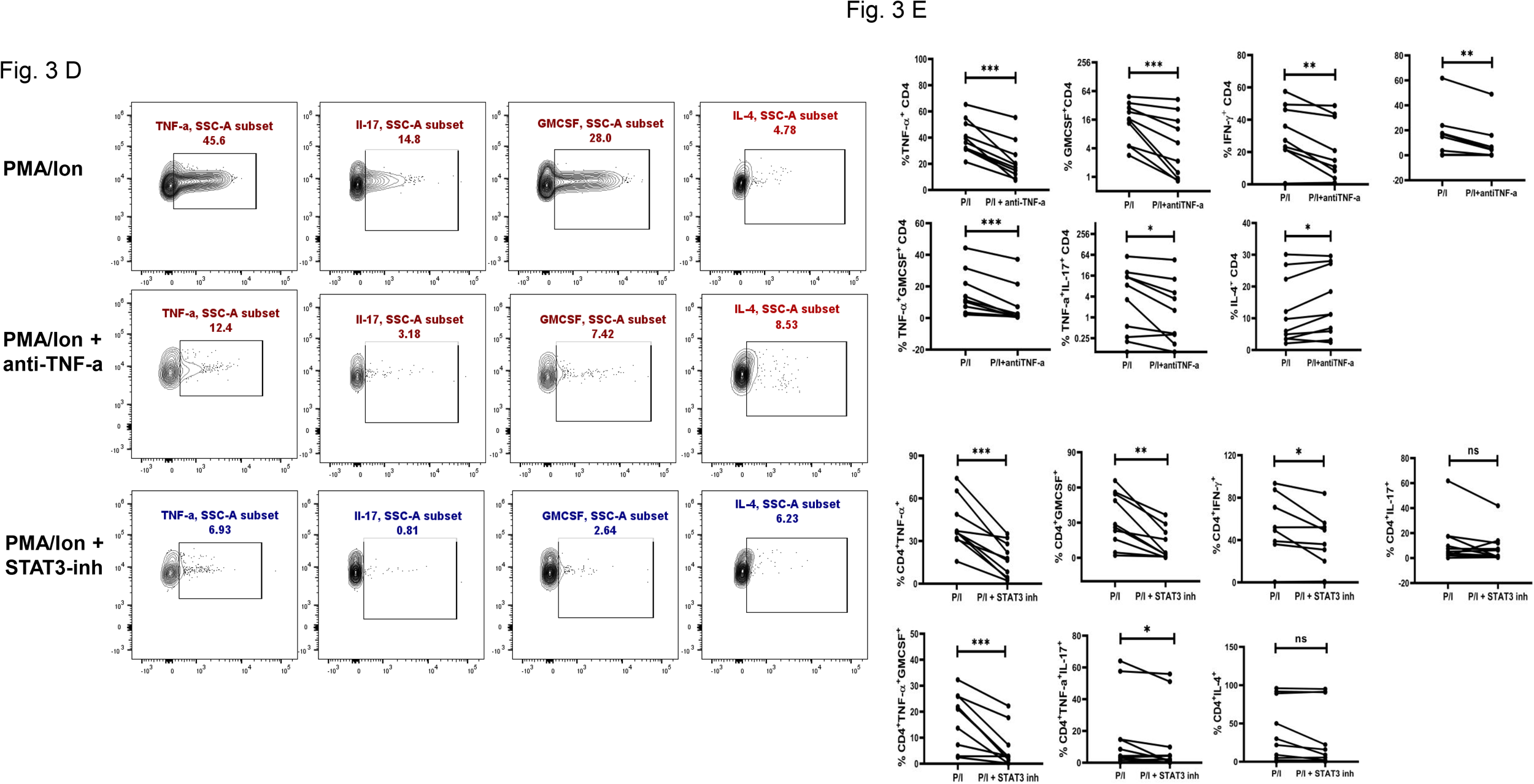
Confocal images depict co-localization of pSTAT1 and pSTAT3 in CD4^+^ T-cells isolated from T2DM patients as compared to healthy controls **(A)**. Expression of pSTAT subsets (STAT1, 3, 4, 5 and 6) in CD4^+^ T cells of T2DM patients following activation with PMA/Ionomycin, represented via multiple overlay histogram plots **(B)**. Increased levels of pSTAT1 and pSTAT3 is observed in T helper cells of T2DM vs healthy controls **(C)**. TNFa and STAT-3 modulates expression of inflammatory cytokines in CD4^+^ T cells isolated from T2DM patients. Pro-inflammatory cytokines levels, including IL-17, IFN-g, and GM-CSF is significantly downregulated along with increase in T2DM CD4^+^T cells upon PMA/Ionomycin stimulation. Similarly, downregulation of TNF-a, IFN-g, and GM-CSF was observed with STAT3 inhibitor treatment independently **(D)**. Paired t test showing decrease in pro-inflammatory cytokines, including IL-17, IFN-g, GM-CSF and aberrant TNF-α^+^GM-CSF^+^ and TNF-α^+^IL-17^+^ secretion upon treatment with a-TNFa and STAT3 inhibitor respectively in CD4+ T cells of T2DM patients **(E)**.

Previous studies have suggested that TNF-α expression may be associated with altered pSTAT3 levels in other chronic autoimmune conditions, such as rheumatoid arthritis [19]. Therefore, we conducted a neutralization assay using a TNF-α monoclonal antibody during activation of CD4^+^ T cells from T2DM patients. Following TNF-α inhibition, we observed a significant decrease in pro-inflammatory cytokines, including IL-17, IFN-γ, and GM-CSF. Additionally, the aberrant TNF-α^+^GM-CSF^+^ and TNF-α^+^IL-17^+^ secretion was also reduced significantly. Among anti-inflammatory cytokines, there was a significant increase in IL-4^+^ cells, while IL-5 and IL-10 levels remained unchanged upon TNF-α inhibition (Fig- 3. D, E).

We also evaluated the effect of pSTAT3 inhibition on cytokine expression wherein treatment with a pSTAT3 inhibitor resulted in a significant decrease in TNF-α IFN-γ and GM-CSF, as well as in the aberrant TNF-α^+^GM-CSF^+^ and TNF-α^+^IL-17^+^ double-positive populations. However, there were no significant changes observed in IL-17, IL-4, IL-5, or IL-10 levels following pSTAT3 inhibition (Fig- 3. F, G).

## Conclusion

The organo pathogenesis of T2DM in the context of inflammation and its cellular sources remain inadequately understood and is primarily defined more by glucose and lipid parameters [20]. This study sought to integrate and analyze meta-inflammation along with biochemical markers. We establish that CD4^+^ T-cells are the primary cells driving meta-inflammation via inflammatory mediators such as TNF-α in T2DM patients through STAT3 and provide a comprehensive characterization of chronic low-grade inflammation in T2DM patients.

The demographic and clinical profile in **Table 1** provides a comprehensive overview of the study population, highlighting key differences in metabolic and inflammatory markers between T2DM patients and healthy controls. The elevated HbA1c, blood glucose, and triglyceride levels in T2DM patients reflect typical metabolic dysregulation associated with diabetes. These parameters, combined with information on lipid profile, renal function, and medication use, lay the foundation for exploring the role of immune dysregulation and inflammatory processes in T2DM.

Our Bio-Plex data demonstrated that approximately 50% of the analytes tested were significantly elevated in T2DM patients compared to healthy controls. These elevated proteins were stratified into high, medium, and low producers based on their secretion levels relative to controls. Notably, multiple pro-inflammatory cytokines, including TNF-α, IL-17, and GM-CSF, exhibited statistically significant elevation, suggesting a complex network of cytokine-driven immune dysregulation mediates meta-inflammation in T2DM as also shown in multiple reports [21, 22].

Not surprisingly, higher levels of anti-inflammatory cytokines viz. IL-4, IL-5 and IL-10 suggested a counter to meta inflammation not mediated through the inflamed T helper cells observed in T2DM. Thus our studies demonstrate while T cell were refractory to anti-inflammatory cytokines, B cells were indeed responsive. This observation was corroborated by antibody profiling, which revealed significantly higher levels of IgA, IgG1, IgG2, IgG3, IgG4, IgE and IgM in T2DM patients, implicating heightened B-cell and plasma cell presence, their inadvertent activation and activity. Persistent cytokine elevation likely contributes to this dysregulation, promoting a feedback loop where innate immune cells, such as macrophages and neutrophils, secrete cytokines including IL-6, IL-1β, IL-15, and IL-8, driving chronic low-grade inflammation.

These findings align with emerging evidence suggesting that meta-inflammation may arise from aberrant interactions between innate immune cells and T helper cells [23, 24], with the latter potentially serving as key regulators in initiating and sustaining this inflammatory cascade [25, 26].

Among the pro-inflammatory cytokines, TNF-α emerged as a central player in the inflammatory response in T2DM patients, consistent with its established role in promoting insulin resistance and contributing to diabetic complications such as nephropathy, neuropathy, and retinopathy [27-29]. Flow cytometry analysis indicated CD3^+^CD4^+^ T helper cells were the primary source of pro-inflammatory cytokines in PBMCs from T2DM patients, including TNF-α. Notably, even in “resting” T-cells, cytokines such as TNF-α, GM-CSF, and IL-17 were secreted without stimulation, suggesting an “activated” CD4^+^ T-cell population that perpetuates inflammation.

TNF-α secretion from effector T-cells appeared a non-specific flare up, while GM-CSF and IL-17 production by memory T-cells suggested a non-specific recall response, potentially directed toward self-antigens.

This study also highlights a significant association between dysregulated biochemical parameters, particularly elevated HbA1c levels, and pro-inflammatory cytokine profiles in T2DM patients. Across three distinct groups categorized by HbA1c levels—DM1 (6.0-7.5%), DM2 (7.5-9%), and DM3 (>9%)—a progressive increase in cytokine signatures, including IFN-γ, TNF-α, and IL-17, was observed. These findings suggest even low level chronic hyperglycaemia exacerbates systemic inflammation by upregulating pro-inflammatory cytokine production. It also reinforces the importance of stringent glycaemic control to mitigate immune dysregulation and chronic low-grade inflammation, which are hallmarks of T2DM pathophysiology.

These results align with previous studies suggesting that sustained T-cell activation and cytokine production are critical drivers of systemic inflammation and auto-inflammatory processes in metabolic disorders [30, 31]. Further research is warranted to elucidate the precise mechanisms underlying these immune perturbations and their potential therapeutic implications.

The activated CD4^+^ T-cell population demonstrated an exaggerated cytokine response, marked by TNF-α, GM-CSF, IL-17, and IFN-γ, which may most probably contribute to organ damage. Both effector and memory T-cells showed increased cytokine responses, reflecting underlying flare-ups and recall reactions, accompanied by notably low levels of anti-inflammatory cytokines, particularly IL-10. The elevated cytokine levels in T-cell compartments were significantly higher than those in healthy controls, suggesting that dysregulated glucose metabolism may alter baseline T-cell cytokine profiles, potentially driving excessive inflammation during infections. While IL-4 and IL-5 responses varied, and seemed to be refractory in T cells, we hypothesize a potential unproductive antibody production role through B cells and a protracted role in wound healing. These findings require further investigation in a larger cohort to clarify their role in T2DM. Thus, our findings indicate that in T2DM, CD4^+^ T-cells exhibit cytokine dysregulation, contributing to meta-inflammation.

TNF-α a pleiotropic cytokine, secreted by multiple sources exhibits pleiotropic effects across various cell types and plays a key role in regulating cellular function and inflammation. TNF-α can signal through TNFRI, which is widely expressed, and TNFRII, which is mostly limited to immune cells. TNF-α signaling via TNFRI induces inflammation and tissue degradation by activating NF-κB and MAPK pathways, which subsequently activate STAT1 and STAT3 [32]. TNFRII, in contrast, is associated with cellular survival and proliferation via STAT5 activation [33].

Mechanistic analyses in our study revealed elevated expression of phosphorylated STAT1 and STAT3 in T2DM patients, where these phosphoproteins were also implicated in the pathogenesis of autoimmune diseases such as rheumatoid arthritis, systemic lupus erythematosus and psoriasis. Confocal microscopy confirmed significantly higher nuclear co-localization of phosphorylated STAT1 and STAT3, further linking these pathways to the pro-inflammatory cytokine response in T2DM. Cytokines such as IFN-γ, IL-6, IL-18, IL-1β along with TNF-α and IL-17 are reported to be increased in pathology with concomitant activation of STAT1 and STAT3, with a majority being activated by TNFRI. TNFRI levels, as shown by Bio-Plex analysis was also found to be elevated because of TNF-α, a complementary mechanism of the immune system to sequester increased levels of TNF-α. These findings support the critical role of STAT3 in TNFRI mediated inflammation and tissue degradation.

Inhibiting TNF-α resulted in a promising reduction in pro-inflammatory cytokines, including GM-CSF, IL-17, and IFN-γ, and decreased the occurrence of TNF-α^+^GM-CSF^+^ and TNF-α^+^IL-17^+^ cells. Additionally, TNF-α inhibition enhanced anti-inflammatory cytokine responses, such as IL-4 secretion. Anti-GM-CSF and STAT5 inhibitors did not significantly affect inflammation, suggesting the primary involvement of the STAT1 and STAT3 pathways. While STAT1 inhibition yielded mixed results without statistical significance, STAT3 inhibition produced similar reductions in pro-inflammatory cytokines as TNF-α inhibition, though it did not show any change with anti-inflammatory cytokine secretion, possibly due to STAT3’s role in IL-10 regulation. A large study cohort might be needed to better understand the role of phospho STAT’s in driving inflammation in these patients

A few limitations of the study include disparity in the age gap between T2DM patients and healthy controls. The male-to-female ratio differs slightly between groups, with a higher percentage of males in the T2DM group. This difference could affect outcomes, as there are known sex-based variations in immune response and metabolic regulation. Most T2DM patients are managed with oral hypoglycaemic agents, with a smaller percentage on insulin. However our findings clearly indicate that glucose or lipid dysregulation control alone through medication was not the best indicator of good immune homeostasis. Clearly immune monitoring wherein medication use directed to regulating immune responses, inflammatory markers, etc. is critically lacking. Ongoing research in our laboratory aims to further characterize the unique T-helper phenotypes, signaling alterations in T2DM, reduction in inflammatory markers with treatment, etc. over a year with a 90-120 day interval.

## Data availability statement

The original contributions presented in the study are included in the article. Further inquiries can be directed to the corresponding authors.

## Ethics statement

The studies involving humans were approved by the Institutional Human Ethics Committee, Institute of Life Sciences (HEC Ref No.: 35/HEC/14). The studies were conducted in accordance with the local legislation and institutional requirements. The participants provided their written informed consent to participate in this study

## Acknowledgments

We would like to thank Dr. Gargee Bhattacharya for her assistance and valuable suggestions in flow cytometry and confocal imaging experiments. We would also like to thank Mr. Paritosh Nath, technical assistant in flow cytometry facility, Ms. Subhranjali Barik for sample collection and Mr. Bhabani Sankar Sahoo, technical assistant in Confocal Imaging facility for their assistance

## Author Contributions

Experimental Design and Conceptualization: SD, SKS. Sample Collection, Processing and Clinical Scoring: DM, RS and PKB. Flow Cytometry Experiments: SKS and RJ. Cytokine Multiplexing: SKS, SS and RJ. Confocal experiment Design, Data Collection and analysis: SD, RJ and SKS.

